# Uptake of health insurance in Malawi in 2019-2020: Evidence from the Multiple Indicator Cluster Survey

**DOI:** 10.1101/2022.08.18.22278931

**Authors:** Wingston Felix Ng’ambi, Farai Chigaru, Takondwa Mwase, Agnes Jack Banda, Joseph Mfutso-Bengo

## Abstract

**Introduction:** Although countries in sub-Sahara Africa (SSA) show progress in implementing various forms of health insurance, there is a dearth of information regarding health insurance in settings like Malawi. Therefore, we conducted this study to determine the uptake of health insurance and describe some of the factors associated with the prevailing uptake of health insurance in Malawi using the 2019-20 Multiple Indicator Cluster Survey (MICS).

**Methods:** This was a secondary analysis of the 2019-20 MICS data. Data were analysed using frequencies and weighted percentages in Stata v.17. Furthermore, since the number of persons with health insurance is very small, we were unable to perform multivariate analysis.

**Results:** A total of 205 (1%) of the 31259 had health insurance in Malawi in 2019-20. Of the 205 individuals that owned health insurance, 118 (47%) had health insurance through their employers while 39 (16%) had health insurance through mutual health organization or ccommunity-based. Men had higher uptake of health insurance than the women. The residents from urban areas were more likely to have a health insurance than those in the rural areas. Persons with media exposure were more likely to own health insurance as compared to their counterparts. There was increasing trend in the uptake of health insurance by wealth of the individual with the poorest being less likely to have health insurance compared to the richest. The persons with no education being least likely to have a health insurance while those with tertiary education were most likely to have health insurance.

**Conclusion:** The uptake of health insurance in Malawi was extremely low. In order to increase the uptake of health insurance, there is need to increase insurance coverage amongst those in formal employment, consider minimizing the geographic, economic and demographic barriers in accessing the health insurance.

## INTRODUCTION

As one of the strategies aimed at achieving the Universal Health Coverage (UHC) by the year 2030, many sub-Saharan African (SSA) countries are at various levels of implementing health insurance schemes in order to improve access to healthcare for their population [1]. The UHC is a key health priority in Sustainable Development Goals. The UHC aims to ensure that everyone has access to high-quality healthcare services that they need, without the risk of financial ruin or impoverishment [2]. Low and middle-income countries (LMICs) in sub-Saharan Africa (SSA) are increasingly turning to public contributory health insurance as a mechanism for removing financial barriers to access and extending financial risk protection to the population [3]. Many countries in Africa are also implementing health Insurance (HI) [4]. For example, Ghana, Kenya, Mali, Nigeria, Rwanda, South Africa, Tanzania, and Zimbabwe have implemented health insurance schemes that seek to improve access to healthcare for their populace [1] [5] [6].

Over the past decade, health insurance uptake has been expanding in SSA [7]. This takes several forms from contributory to community-based Health Insurance (CBHI). The World Health Organisation (WHO) observed that the high cost through direct out-of-pocket health expenditure is a major barrier to the achievement of UHC which is a target of the SDG [8]. Studies have shown that health insurance coverage is a key factor in accelerating progress towards the achievement of UHC [1]. The Malawi Ministry of Health made tremendous progress towards HI by doing several working papers and conducting assessment on the feasibility of the SHI in Malawi. The key finding of the assessment by Gheorghe et al is that HI is feasible in Malawi and that the National Health Insurance Scheme’s (NHIS) is a way of generating additional health revenue [4]. Several studies in SSA have shown limited access to health insurance [1] [9]. Furthermore, there have been very few studies that assess the uptake of health insurance in Malawi. Therefore, we conducted this study to determine the uptake of health insurance and describe some of the factors associated with the prevailing uptake of health insurance in Malawi using the 2019-20 Multiple Indicator Cluster Survey (MICS).

## METHODS

### Study Design

The study used data from the Multiple Indicator Cluster Survey 2019-2020, Round 6. The MICS conducted nationally representative surveys in Malawi between 2006 and 2019. The MICS surveys measure key indicators that allow countries to generate data for use in policies, programmes, and national development plans, and to monitor progress towards the Sustainable Development Goals (SDGs) and other internationally agreed upon commitments [10].

### Data collection procedure

The surveys’ data collection technique included using a standard questionnaire that is equivalent across nations [11]. The questionnaire is frequently translated into the major local languages of the participating countries. The translated questionnaires along with the English-language version, are pretested in English and the local dialect to guarantee their validity. The details of the sampling methodology, procedures, and implementation can be found in the report by MICS 2019-20 [10].

### Sampling procedure and size

The sampling procedure employed in the surveys involved a two-stage stratified sampling procedure, where Malawi was grouped into urban and rural areas. The first stage involved the selection of clusters usually called enumeration areas (EAs) and the second stage consisted of the selection of a household for the survey [10]. In this study we included both men and women that were interviewed in the MICS round 6 in Malawi.

### Study variables

The outcome variable of this study was health insurance coverage. This was derived from the question “are you covered with any health insurance?”. The response is coded as 0 = “No” and 1 = “Yes”. The explanatory variables were age, wealth status, level of education, marital status, frequency of reading newspaper or magazine, frequency of listening to the radio, and frequency of watching television. Age was recoded as 15–19, 20–24, 25–29, 30–34, 35–39, 40–44, 45–49, 50–54, 55–59, 60–64. Wealth status was categorized as poorest, poorer, middle, richer, and richest. Education was classified into four categories: no education, primary education, secondary education, and higher education. The frequency of reading newspaper or magazine, frequency of listening to radio, and frequency of watching television were respectively captured as not at all, less than once a week, at least once a week, and almost every day. Our study variables were based on previous literature [1] [2] [12].

### Statistical analysis

Data were analysed in STATA 17.0 (Stata Corp., College Station, TX). We calculated frequencies and percentages. Analysis incorporated the weights since data were from complex survey design. Furthermore, since the number of persons with health insurance is very small then we were unable to perform multivariate analysis. The STrengthening the Reporting of OBservational studies in Epidemiology (STROBE) guidelines were used to conduct and report on the findings of this study [13].

### Ethical Consideration

The individual consent was conducted by National Statistical Office (NSO) of Malawi during the MICS 2019-20 round 6. We obtained permission to use this data from the UNICEF MICS. The Malawi dataset is freely available for download from https://mics.unicef.org/surveys (accessed on 31 March 2022).

## RESULTS

### Characteristics of the men and women interviewed in Malawi in 2019-20

The characteristics of the men and women assessed for health insurance coverage are shown in Table 1. A total of 31259 men and women were assessed. Of these; 24494 (78%) were females and 26138 (84%) were from rural areas. The majority of the people were aged 15-19 years (22% of 31259) while the least were aged 45-49 years (7% of 31259). The majority of the persons (63% of 31259) had primary education while the minority had no education (0% of 31259). There was increasing trend in the proportion of individuals by wealth index quintile (see Table 1). There was variation in exposure to media with the majority having access to information through the radio (56% of 31259) while the minority accessed information through internet (9% of 31259). Seven percent of the individuals did not indicate their levels of education.

**Table 1:** Characteristics of persons evaluated for health insurance coverage in Malawi in 2019-20.

| Characteristics | n | % |
| --- | --- | --- |
| **Total** | 31259 | 100.0 |
| **Sex** |  |  |
| *Male* | 6765 | 21.6 |
| *Female* | 24494 | 78.4 |
| **Location** |  |  |
| *Urban* | 5121 | 16.4 |
| *Rural* | 26138 | 83.6 |
| **Age group** |  |  |
| 15-19 | 7639 | 21.6 |
| 20-24 | 5981 | 18.8 |
| 25-29 | 4778 | 17.5 |
| 30-34 | 4023 | 12.9 |
| 35-39 | 3779 | 12.1 |
| *40-44* | 2895 | 9.3 |
| *45-49* | 2164 | 6.9 |
| **Highest education level** |  |  |
| *No education* | 15 | 0.0 |
| *Primary* | 19604 | 62.7 |
| *Secondary* | 8499 | 27.2 |
| *Tertiary* | 883 | 2.8 |
| *Missing* | 2258 | 7.2 |
| **Wealth index quintile** |  |  |
| *Poorest* | 5565 | 17.8 |
| *Poorer* | 5670 | 18.1 |
| *Middle* | 5862 | 18.8 |
| *Richer* | 6604 | 21.1 |
| *Richest* | 7558 | 24.2 |
| **Media exposure** |  |  |
| ***Television*** |  |  |
| *No* | 23785 | 76.1 |
| *Yes* | 7474 | 23.9 |
| ***Radio*** |  |  |
| *No* | 13904 | 44.5 |
| *Yes* | 17355 | 55.5 |
| ***Newspaper*** |  |  |
| *No* | 25133 | 80.4 |
| *Yes* | 6126 | 19.6 |
| ***Internet*** |  |  |
| *No* | 28448 | 91.0 |
| *Yes* | 2811 | 9.0 |

### Uptake of health insurance in Malawi in 2019-20

The uptake of health insurance in Malawi in 2019 is shown in Table 2. A total of 205 (1%) of the 31259 had health insurance in Malawi in 2019. There were variations in the uptake of uptake of health insurance by sex, age, rural/urban location, wealth quintile and media exposure. Men had higher uptake of health insurance than the women. The residents from urban areas were more likely to have a health insurance than those in the rural areas. Persons with media exposure were more likely to have access health insurance as compared to their counterparts (see Table 2). There was increasing trend in the uptake of health insurance by wealth of the individual with the poorest being less likely to have health insurance compared to the richest. Furthermore, the persons with the no education being least likely to have a health insurance while those with tertiary education are most likely to have health insurance.

**Table 2:** Distribution of coverage of health insurance by socio-demographic characteristics in Malawi in 2019-20.

| Characteristics (n= 31259) | Has health insurance |  |  |  |
| --- | --- | --- | --- | --- |
|  | No |  | Yes |  |
|  | N | % | n | % |
| **Total** | 31054 | 99.2 | 205 | 0.8 |
| **Sex** |  |  |  |  |
| *Male* | 6705 | 99.0 | 60 | 1.0 |
| *Female* | 24349 | 99.3 | 145 | 0.7 |
| **Location** |  |  |  |  |
| *Urban* | 4987 | 97.2 | 134 | 2.8 |
| *Rural* | 26067 | 99.7 | 71 | 0.3 |
| **Age group** |  |  |  |  |
| *15-19* | 7618 | 99.5 | 21 | 0.5 |
| *20-24* | 5962 | 99.6 | 19 | 0.4 |
| *25-29* | 4752 | 99.5 | 26 | 0.5 |
| *30-34* | 3976 | 99.0 | 47 | 1.0 |
| *35-39* | 3742 | 99.0 | 37 | 1.0 |
| *40-44* | 2859 | 98.5 | 36 | 1.5 |
| *45-49* | 2145 | 98.7 | 19 | 1.3 |
| **Highest education level** |  |  |  |  |
| *No education* | 15 | 100.0 | 0 | 0.0 |
| *Primary* | 19584 | 99.9 | 20 | 0.1 |
| *Secondary* | 8430 | 99.0 | 69 | 1.0 |
| *Tertiary* | 769 | 87.0 | 114 | 13.0 |
| **Wealth index quintile** |  |  |  |  |
| *Poorest* | 5564 | 100.0 | 1 | 0.0 |
| *Poorer* | 5664 | 99.9 | 6 | 0.1 |
| *Middle* | 5855 | 99.8 | 7 | 0.2 |
| *Richer* | 6576 | 99.6 | 28 | 0.4 |
| *Richest* | 7395 | 97.2 | 163 | 2.8 |
| **Media exposure** |  |  |  |  |
| ***Television*** |  |  |  |  |
| *No* | 23742 | 99.8 | 43 | 0.2 |
| *Yes* | 7312 | 97.4 | 162 | 2.6 |
| ***Radio*** |  |  |  |  |
| *No* | 13859 | 99.6 | 45 | 0.4 |
| *Yes* | 17195 | 99.0 | 160 | 1.0 |
| ***Newspaper*** |  |  |  |  |
| *No* | 25059 | 99.7 | 74 | 0.3 |
| *Yes* | 5995 | 97.6 | 131 | 2.4 |
| **Internet** |  |  |  |  |
| *No* | 28309 | 99.5 | 139 | 0.5 |
| *Yes* | 2745 | 97.2 | 66 | 2.8 |

### Type of health insurance accessed by Malawians in 2019-20

The types of health insurance owned by Malawians by 2019 are shown in Table 3. Of the 205 individuals that had health insurance, 118 (47%) had health insurance through their employers while 39 (16%) had health insurance through mutual health organization or community-based schemes. Nineteen percent of the persons had health insurance through social security (see Table 3).

**Table 3:** Types of health insurance accessed by Malawians in 2019-20.

| **Type of health insurance (n=250)** | **n** | **%** |
| --- | --- | --- |
| Mutual health organization/Community-based | 39 | 15.6 |
| Through employer | 118 | 47.2 |
| Purchased privately | 46 | 18.4 |
| Social security | 47 | 18.8 |

### Health insurance coverage across the districts of Malawi in 2019-20

The coverage of health insurance by district in 2019 in Malawi is shown in Table 4. Zomba (1.3%), Thyolo (1.4%), Mzimba (1.7%) and Blantyre (2.5%) were the districts with the highest uptake of health insurance. Chitipa (0%) and Ntchitsi (<0.01%) were the two districts with nearly no health insurance coverage. Zomba (9%), Mzimba (5%) and Kasungu (5%) were the three districts with the highest uptake of health insurance in the urban areas. Blantyre and Thyolo were the two districts with the highest health insurance coverage (2%) in rural areas.

**Table 4:** Coverage of health insurance by district and rural-urban locations in Malawi in 2019-20.

| **District** | **Overall (n=31259)** | **Rural (n=26138)** | **Urban (n=5121)** |
| --- | --- | --- | --- |
| Balaka | 0.1 | 0.0 | 1.4 |
| Blantyre | 2.5 | 1.6 | 3.1 |
| Chikwawa | 0.5 | 0.6 | 0.0 |
| Chiradzu | 0.7 | 0.6 | 4.0 |
| Chitipa | 0.0 | 0.0 | 0.0 |
| Dedza | 0.1 | 0.1 | 0.0 |
| Dowa | 0.3 | 0.3 | 0.0 |
| Karonga | 0.6 | 0.2 | 2.0 |
| Kasungu | 0.9 | 0.1 | 5.0 |
| Likoma | 0.7 | 0.5 | 2.4 |
| Lilongwe | 0.8 | 0.0 | 2.0 |
| Machinga | 0.1 | 0.1 | 2.7 |
| Mangochi | 0.1 | 0.0 | 1.2 |
| Mchinji | 0.3 | 0.2 | 0.6 |
| Mulanje | 0.4 | 0.3 | 4.4 |
| Mwanza | 0.8 | 0.6 | 1.5 |
| Mzimba | 1.4 | 0.1 | 5.1 |
| Neno | 0.4 | 0.4 | 0.0 |
| Nkhata B | 0.1 | 0.1 | 1.0 |
| Nkhotako | 0.4 | 0.3 | 0.9 |
| Nsanje | 0.6 | 0.1 | 3.6 |
| Ntcheu | 0.1 | 0.1 | 0.0 |
| Ntchisi | 0.0 | 0.0 | 1.6 |
| Phalombe | 0.1 | 0.1 | 1.7 |
| Rumphi | 0.1 | 0.1 | 0.0 |
| Salima | 0.5 | 0.2 | 2.9 |
| Thyolo | 1.7 | 1.8 | 0.0 |
| Zomba | 1.3 | 0.3 | 8.7 |

## DISCUSSION

This is the first analysis using data from Malawi Indicator Cluster Survey to focus on the coverage of health insurance in Malawi and contributes hugely in understanding uptake of health insurance in a predominantly free public health care system. The following were the key findings: the coverage of health insurance was extremely low; urban population had better health insurance coverage than the rural population; health insurance varied by age and sex as well as education and wealth; the minority of the those with insurance accessed it through private purchase or community insurance; and exposure to media was associated with more uptake of health insurance.

Our study found extremely low coverage of health insurance in Malawi. This is a cause of concern as far as achievement of UHC. However, other studies have bemoaned low coverage of health insurance in similar settings. For example; the level of health insurance coverage in SSA is low; only 8 of the 36 countries examined had a mean level of insurance coverage with any type of health insurance of above 10%, while only 4 had a coverage level of above 20% [2]. The coverage in the other settings have been higher than what was reported in Malawi [14]. The differences may be attributed to the differences in the socio-demographic characteristics of the individuals as well as different funding mechanisms for the National Health Insurance systems. Further explanation for the observed difference in the uptake of health insurance in Malawi compared to the other countries is that in Malawi healthcare in the public sector (the largest provider of health services) is provided to the populace free of charge where as in the other countries the populace pay for most healthcare services.

Similar to our study, exposure to media, socioeconomic rank and the level of education had the greatest contribution to inequality in coverage with any type of health insurance in SSA [2]. A study conducted in Nepal, Ghana and Ethiopia also found that mass media exposure was significantly associated with high enrollment in HI [15] [16] [17] [18]. This has implications in utilization of the various media communication channels in making strategic communication plans to promote health insurance uptake in Malawi or any other similar settings. A study in SSA by Barasa et al found that wealthier individuals were more likely to have a health insurance compared to those with less income [2]. Generally, the higher an individual’s wealth level is, the more likelihood he will participate in a health care program in SSA [2] [19] [20] and this is similar to what was observed in China, Spain and USA [18] [21] [22] [23] [24]. The implication of the association between wealth and insurance uptake for settings like Malawi would be to consider a minimal amount of health insurance schemes for the less privileged since the less privileged are the most marginalized in uptake of health insurance. Persons with low education level have lower uptake of health insurance as observed in other similar settings like Kenya [20] [24] [25]. Level of education proxies literacy level therefore people with little knowledge and understanding on universal health care have low use of health insurance [20]. For health insurance uptake to improve then people should be well informed on the same hence literacy in the health systems of a country should be undertaken [20].

There are variations in the uptake of health insurance by gender. Whereas some studies like the one done in Kenya [25] did not find any significant difference in health insurance with gender although some studies have indicated lower insurance uptake by men than women due to their greater need for more healthcare services [25]. The lower participation of women in health insurance in Malawi may be attributed to the low economic and social position of women relative to men as observed in Ghana [26]. Women are more economically-disadvantaged and usually have lower access to interventions and programs due to the highly patriarchal nature of most rural communities.

Our study shows that the urban residents had more uptake of health insurance compared to their rural counterparts. Amu et al. examined the variations in health insurance coverage in four countries (Ghana, Kenya, Nigeria, and Tanzania) which were the first SSA countries to launch developmental plans in the early 1960s [27]. Amu et al. realized that in most of the countries, the probability of being covered by health insurance was lower among urban dwellers [27]. However, a study conducted in the USA is consistent with our study as it also shows higher uptake of health insurance uptake in the urban settings than in the rural settings [28]. The findings from Amu et al. are a contradiction to what we have observed in Malawi.

Based on the findings from this study, the minority of the those with insurance accessed it through private purchase or community insurance. The uptake of these types of insurance provides an opportunity for community-based health insurance schemes that have proven to be key in bridging the insurance gaps in other similar settings. One of the key strategies to be used to increase the uptake of community-based health insurance (CBHI) in setting like Malawi include increased awareness [7] [29]. Although health insurance from social security was available in Malawi for some persons, such health insurance may not be sustainable especially if they depend on donor funding. Hence the quest to go for CBHI in settings like Malawi.

The strength of the study is utilization of a nationwide survey data for 2019-20 MICS. One weakness of the paper is absence of some key variables like severity of diseases or the diseases suffered by the individuals in Malawi. These variables have been reported in other settings as being key drivers of uptake of health insurance [30]. The other limitation of this study is that the numbers of individuals that had a health insurance in Malawi was very low hence we could not perform a multivariate analysis.

## CONCLUSION

The uptake of health insurance in Malawi is extremely low. Furthermore, our findings also show that health insurance in Malawi is highly inequitable. In order to achieve UHC by the year 2030, many sub-Saharan African (SSA) countries need to implement equitable health insurance schemes that seek to improve access to healthcare for their population. We recommend increased public education on the benefits of being covered by health insurance using the mass media which we found to be an important factor associated with health insurance coverage.

The focus of such mass media education could target the less educated rural dwellers, males, the poor, and the young.

## Data Availability

Downloaded from Measure DHS

https://mics.unicef.org/surveys

## DECLARATIONS

We declare that there is no conflict of interest in publishing this paper.

## AUTHORS’ CONTRIBUTIONS

WFN led the manuscript writing, conducted data management and analysis; TM, FC & JMB advised on the data analysis and policy insights on the paper. All authors read and approved the final manuscript.

## ACKNOWLEDGEMENT

The authors would like to thank the UNICEF for allowing us to use the Malawi 2019-20 MICS data.

